# Multi-dimensional prediction of suicidality and non-suicidal self-injury transition in children: from general psychopathological, behavioural, and neurobiological perspectives

**DOI:** 10.1101/2022.11.21.22282608

**Authors:** Xue Wen, Qiyang Qu, Yinzhe Wang, Xiaoqian Zhang, Zaixu Cui, Runsen Chen

## Abstract

**IMPORTANCE:** Accurate prediction of suicide or non-suicidal self-injury (NSSI) among children within a uniform time frame is an essential but challenging task. Furthermore, few studies have comprehensively considered clinical, behavioural, and neurobiological factors to produce multi-dimensional prediction models.

**OBJECTIVE:** To examine predictive effects of general psychopathology, behavior inhibition system, and brain signature on children’s suicidality or NSSI transition.

**DESIGN, SETTING, AND PARTICIPANTS:** We adopted a retrospective and longitudinal methodology by utilising the data from the Adolescent Brain Cognitive Development (ABCD) cohort. In total, 9332 individuals aged 9-10 years without any suicidality or non-suicidal self-injury (NSSI) history at baseline were included in our analyses. Then, four subgroups were generated based on whether they had developed suicide ideation (Healthy control [HC]-SI), NSSI (HC-NSSI) or suicide attempt (HC-SA) in a year, while the remaining group was considered a control group (HC-HC).

**MAIN OUTCOMES AND MEASURES:** Participants suicidal behaviors and non-suicidal self-injury behaviors were assessed with the Kiddle Schedule for Affective Disorders and Schizophrenia. Meanwhile, general psychopathology (i.e., *p-factor*) was calculated based on scores of Child Behavior Checklist, behavioral inhibition system (BIS) was assessed though BIS/BAS scale, and the brain morphometrics were also collected though sMRI. Multinomial logistic regression models were used for assessing the predictive effects of general psychopathology, behavioral inhibition system, and whole-brain cortical area on children’s STB and NSSI transition.

**RESULTS:** As a result, we found higher general psychopathology in baseline predicted higher NSSI (1.52 [1.28-1.80]), SI (OR=1.34 [95%CI 1.17-1.53]) and SA (2.05 [1.34-3.14]) risk in a year. From a behavioural perspective, higher BIS sensitivity predicted higher SI (2.05 [1.61, 2.61], and NSSI (1.68 [1.24, 2.28]) in a year. From a neurobiological perspective, abnormalities in the cortical area of the superior insula, inferior frontal area, superior temporal area, and superior precentral area were all shown to be associated with children’s NSSI, SI and SA in the future.

**CONCLUSIONS AND RELEVANCE:** This study is the first to look at the predictive factors for the different transitions of NSSI and suicidal behaviour from the biopsychosocial framework. Our findings offered empirical evidence on the predictive effect of baseline general psychopathology, BIS sensitivity and biological marker on children’s suicidality or NSSI in a year, providing early biomarkers for all types of transition. In this case, the early identification of those factors may facilitate the development of early prevention or intervention that could potentially alleviate more relevant public health issues.

**Key Points:** *Question:* Could general psychopathology, behavior inhibition system, and brain signature predict suicidality or NSSI transition in children?

*Findings:* In a longitudinal observational study (9332 children), higher general psychopathology at baseline predict higher risk of suicidality and NSSI transition in a year. Meanwhile, higher BIS sensitivity also predict higher risk of suicidality and NSSI transition. To note, abnormalities in the cortical area of the superior insula, inferior frontal area, superior temporal area, and superior precentral area were all shown to be associated with children’s suicidality and NSSI transition.

*Meaning:* The early identification of biopsychosocial factors associated with suicidality or NSSI transition in children could facilitate early prevention.

## Introduction

Self-harm, as an umbrella terminology, refers to non-suicidal self-injury (NSSI), suicide ideation or suicidal behaviours (also frequently being referred to as “Suicidal thoughts and behaviour”; STBs), is an alarming public health problem in adolescents worldwide^1^. As a precursor of STBs, it has been reported that the pooled NSSI prevalence was 17% amongst adolescents^2^, sharply increasing to over 40% during acute stress times such as during the pandemic^3^. A population-based study of 82 countries has reported that the prevalence of suicidal ideation was nearly 14.0% among adolescents ^4^. Additionally, as the second leading cause of death for adolescents ^5,6^, suicide behaviour’s prevalence accounted for 11.0% among adolescents in a large cross-national study^7^. The negative effect of NSSI and STBs on adolescents’ development outcomes has been highlighted in several systematic reviews and meta-analyses, including changes in brain structure ^8^, poorer academic outcomes, higher mental health problems, and delinquent behaviour, which may last from childhood to adulthood ^9^.

Therefore, it has become an obligation for health professionals to identify the underlying mechanism that can prospectively predict the occurrence process, which is essential for developing effective prevention or intervention program at an early stage. The “continuum” model has recognized that these self-harm behaviours should best be conceptualized along a continuum, highlighting the existing shared and unique risk factors across these behaviours ^10^. For example, several studies have highlighted the modifiable characteristics associated with NSSI and STBs, including cognitive appraisal factors ^11^, psychiatric morbidity^12^, and lower levels of social support. Nevertheless, to what extent these factors contribute to the process of emergence for these subgroups of self-harm behaviours remains equivocal. More importantly, researchers have long been interested in how people who accidentally persist in NSSI are similar or differ from individuals who report suicidal ideation or engage in suicide attempts, but the nature of the underlying mechanism remained unclear.

The impact of internalizing and externalizing psychopathology on the occurrence of NSSI or STBs has received extensive attention. For example, a recent meta-analysis found that the symptoms of major depressive disorder and anxiety disorders predicted the onset of NSSI^13^. In line with this, Hills and colleagues (2009) found that externalizing psychopathology (e.g., antisocial personality disorder) was the risk factor for the later development of new-onset suicide attempts. Lately, instead of perceiving mental disorders as distinct, episodic and categorical conditions, an increasing number of researchers have suggested that one general psychopathology dimension exists, which is referred to as the *p-factor* ^14^, or general psychopathology^15,16^. The *p-factor* has been suggested to be strongly associated with NSSI and suicidality^17^. However, the prediction role of the *p-factor* on the similar or distinguished trajectory of occurrence of NSSI and STBs still requires further exploration.

Additionally, behavioural inhibition systems (BIS), as an aversive motivational system activated by signals of punishment, have been a major focus of NSSI and STBs studies. Generally speaking, a common assumption suggested by most theories suggests that NSSI or STBs is primarily motivated by the strong desire to escape from intense emotional pain or aversive situations ^18,19^. In this case, an individual’s higher sensitivity to aversive stimuli may lead to the consequence of more maladaptive avoidant behaviours ^20^. This perspective is supported by several studies, which found that individuals’ BIS was positively associated with their likelihood of NSSI or STBs in several cross-sectional studies^21,22^. In addition, a previous study found that individuals with a higher level of BIS reported more severe forms of suicidality^20^. However, to what extent the BIS predicted the onset of different subgroups of NSSI or STBs still to this day remains unclear.

Despite detecting clinical and behavioural predictors, neuroimaging is the most promising new direction for predicting STB and NSSI transition. A recent meta-analysis of 131 neuroimaging studies found structural and functional alterations in the brain associated with STB, which converges in the prefrontal cortex (PFC), insula, cingulate cortex (CC), temporal regions, as well as other cortical regions.^23^ Meanwhile, the same regions were also shown in a recent neuroimaging meta-analysis about NSSI in youth.^24^ Indeed, due to the high co-occurrence of STB and NSSI, it is not surprising to find a common neurobasis underlying these behaviours.^3^ However, there is a scarcity of studies that combine 1) longitudinal study design, 2) large sample sizes, and 3) target subjects in critical periods of development (i.e., children). Emphasis on detecting neurobiological markers of future STB and NSSI transition is necessary due to its importance in guiding our preventive programs.

Using data from a large cohort study and following the framework of the biopsychosocial model, we thus aimed to investigate the risk factor which predicted the NSSI and STBs transition, including neuroimaging, general psychopathology, and impulsivity behaviours perspectives.

## Methods

### Data sources

We included data from the Adolescent Brain and Cognitive Development (ABCD) “Curated Annual Released 4.0 version” ^25^. Data for all included measures were collected at baseline for 11,875 participants between 2016 and 2018, with the excetion of STB and NSSI transition-related data being collected at 1-year follow-up. Written informed consent from caregivers and verbal assent from children were obtained. All procedures were approved by the centralised institutional review board (IRB) from the University of California, San Diego, and each study site obtained approval from its local IRB.

### Measurements

#### STB and NSSI transition

The child-report version of the suicide module from the computerised Kiddle Schedule for Affective Disorders and Schizophrenia (KSADS, Lifetime version) was utilized to assess children’s lifetime SI, NSSI, and SA.^39^ To note, participants were coded as having SI if they endorsed SI in any of the following five domains: passive, active but non-specific, specific method, active with intent, and active with a plan. Both baseline and 1-year follow-up assessments were considered to describe children’s STB and NSSI transition.

#### General Psychopathology

The parent-report version of the Child Behavior Checklist (CBCL for ages 6 to 18) was used to assess children’s problematic behaviours that needed attention^17^. All of the 119 items were scored on a 3-point Likert scale ranging from 0 (“not true”) to 2 (“very true”). According to an established bifactor model, which has been tested for good reliability and validity, four-dimensional psychopathology factors were derived. Only 66 items were concluded in the final model, and then the general factor scores (named *p-factor*) were calculated for each participant. The general psychopathology scores were only calculated at baseline^26^. Due to the parent-child discrepancy observed in KSADS diagnoses in previous studies, two items (i.e., “Deliberately harms self or attempts suicide”, “Talks about killing self “)out of the bi-factor model were selected as the additional STB and NSSI screening items.

#### Behaviour System

The Behavioral Inhibition/Activation Scales (BIS/BAS) administered to children were also utilized in the present study. They were both scored on a 4-point Likert scale ranging from 0 (“not true”) to 3 (“very true”). The BIS scale ^27^, consisting of 7 items, is concerned with motivation to avoid aversive outcomes.

#### Brain Morphometry

All preadolescents underwent structural Magnetic Resonance Imaging (sMRI) according to the ABCD study’s standardised protocol, while the scanning parameters, pre-processing, and analytical pipelines were described in considerable detail.^40,43^ FreeSurfer v5.3 (http://surfer.nmr.mgh.harvard.edu) was used to process the locally acquired T1-weighted images to estimate global cortical area. Then the cerebral cortex was parcellated into 74 regions according to the Desikan-Killiany atlas.^44^ Only scans that passed protocol compliance and quality control were used for corresponding analyses.

### Statistical Analysis

In total, the 11,875 participants were subset to 9332 if they 1)reported no lifetime SI, SA, and NSSI at baseline, 2) passed the CBCL screening item reported by their caregivers (i.e., got a “0” score in both items), and 3) had complete sociodemographic and STB and NSSI transition-related information. Sociodemographic information, including children’s age, sex, race/ethnicity, and data acquisition sites, were used as covariates. All statistical analyses were performed using SPSS (version 26.0) and MATLAB (version 2022a). An overview of our analysis pipeline can be seen in Figure 1, while detailed information related to all the used variables is shown in Appendix p1.

**Figure 1.**
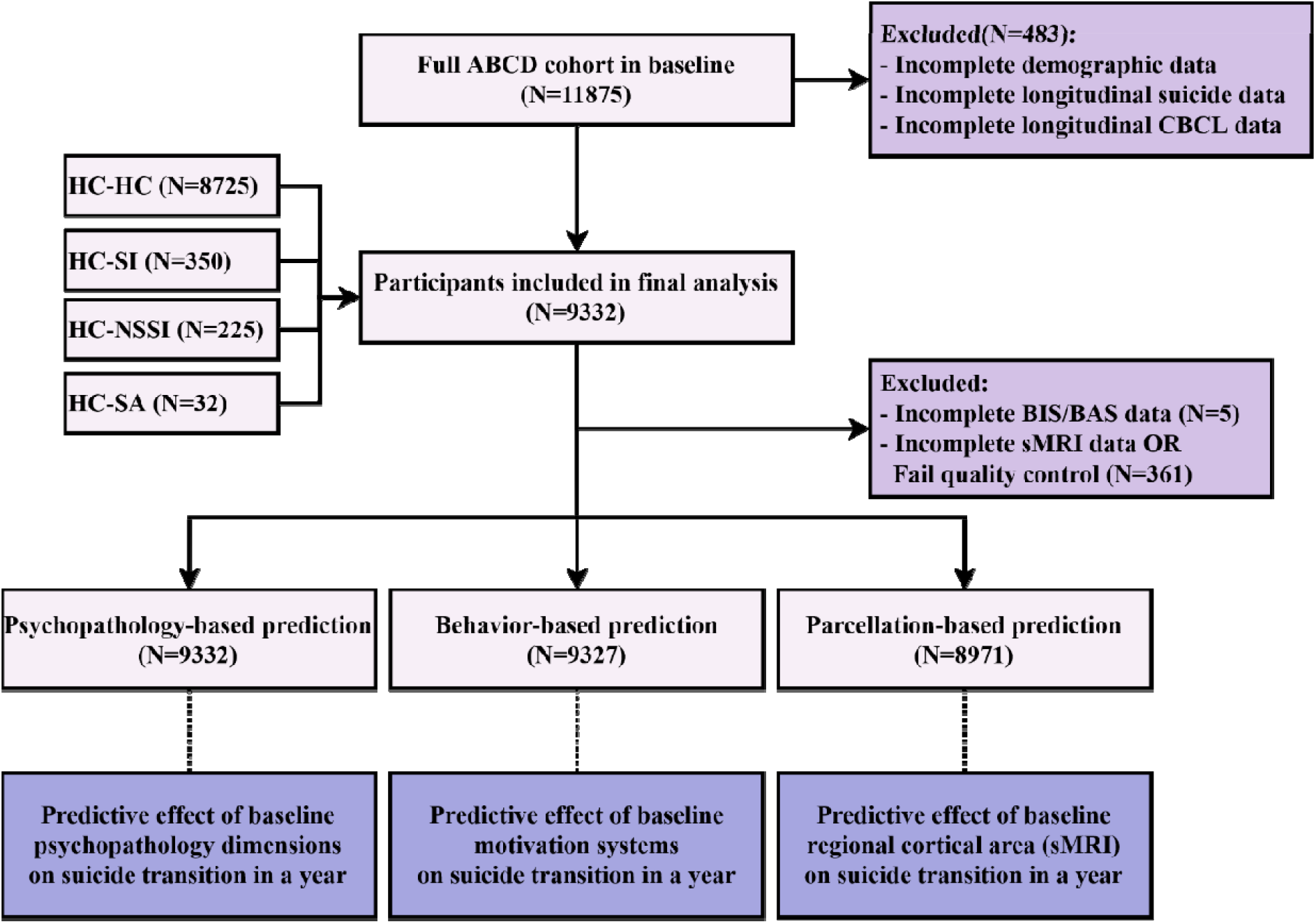
The operational flowchart of the present study.

First, participants were divided into four groups due to their mental health condition in 1-year follow-up. Children were classified as 1) the HC-SA group if they reported SA from baseline and 1-year follow-up, 2) the HC-NSSI group if they had NSSI but no SA, 3) the HC-SI group if they only reported SI rather than self-harm behaviour or 4) the HC-HC group if they reported no lifetime SI, SA, and NSSI themselves and passed the CBCL screening items during that time.

Next, we assessed whether subjects exhibited differences in dimensional psychopathology, behavioural system, or brain morphometry at baseline, even if they did not start to transit. The multivariate analysis of covariance (MANCOVA) was used to compare the group differences within the four subgroups (i.e., HC-HC, HC-SI, HC-NSSI, and HC-SA), and then pairwise comparisons were further conducted.

Finally, the predictive effects of dimensional psychopathology, behavioural system, and brain morphometry on children’s STB and NSSI transition in a year were further examined. A series of multinomial logistic regression models were performed, and an odds ratio (OR) was calculated for each analysis to characterize the strength of the associations. For the psychopathology aspect, the independent variables included participants’ factor scores on general, externalizing, attention-deficit/hyperactivity disorder (ADHD), and internalizing psychopathology. For the behavioural aspect, the independent variables included subscale scores of BIS, BAS-reward, BAS-drive, and BAS-fun seeking. For sMRI analysis, only 8971 scans that passed the quality control were taken into the subsequent analyses. Then, all 74 cortical areas were put into the model for prediction analysis. The false discovery rate (FDR) was further used to avoid the potential bias arising from multiple comparisons.^46^

## Results

For 9332 eligible subjects without any STB and NSSI history at baseline, four subgroups were generated based on whether they developed SI (HC-SI, N=350), NSSI (HC-NSSI, N=225), SA (HC-SA, N=32) in a year or not (HC-HC, N=8725). The baseline demographic details of four subgroups are summarized in Table 1.

### General psychopathology predicts children’s STB and NSSI transition

We found that the children showed differences in general psychopathology even before the transition occurred (*F*=19.77, *P*<0.001). To note, after FDR correction, the HC-SA group performed the highest level of general psychopathology at baseline, while the HC-HC group performed the lowest in general psychopathology compared to the STB and NSSI transition groups (Figure 2). Furthermore, a higher level of general psychopathology was shown to predict severer SI (OR=1.34 [95%CI 1.17-1.53]), NSSI (1.52 [1.28-1.80]), and a higher frequency of SA (2.05 [1.34-3.14]) transition risk in a year, suggesting a common marker for children’s STB and NSSI transition (Figure 3).

**Figure 2.**
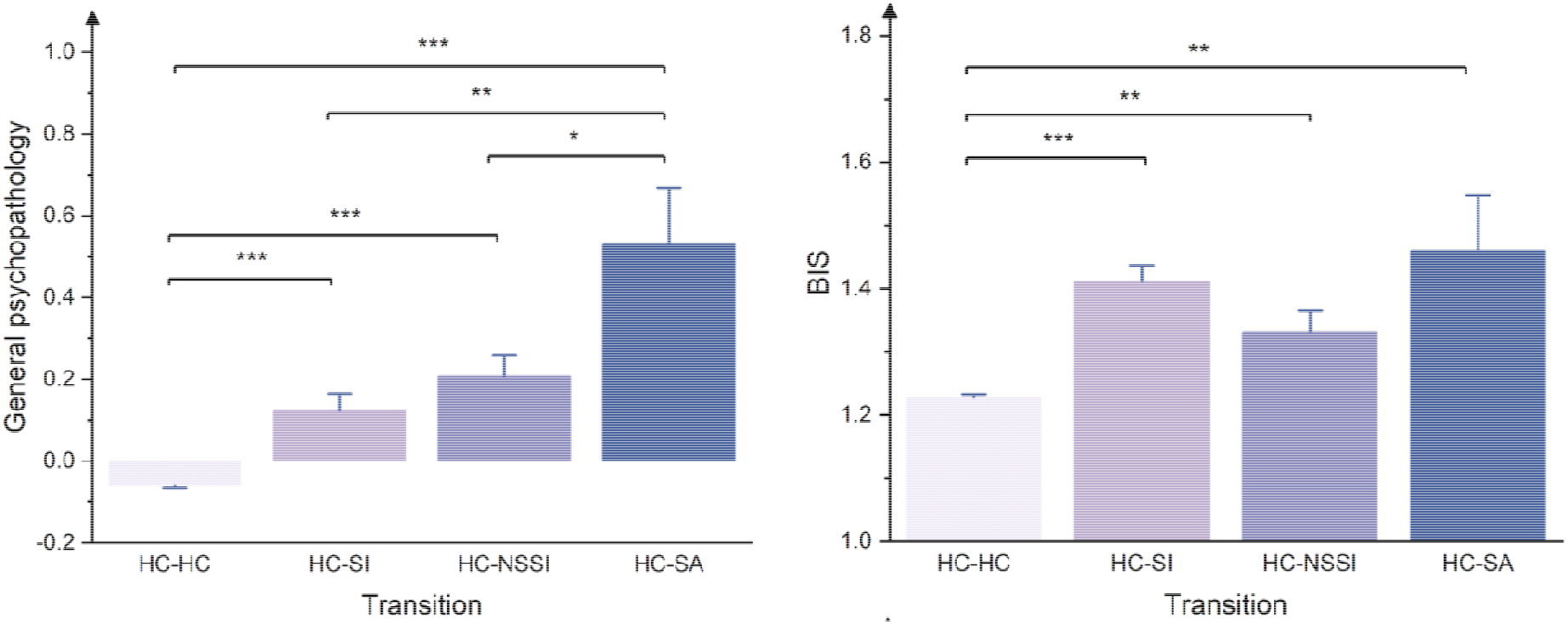
Characteristics of general psychopathology and BIS before STB and NSSI transition

**Figure 3.**
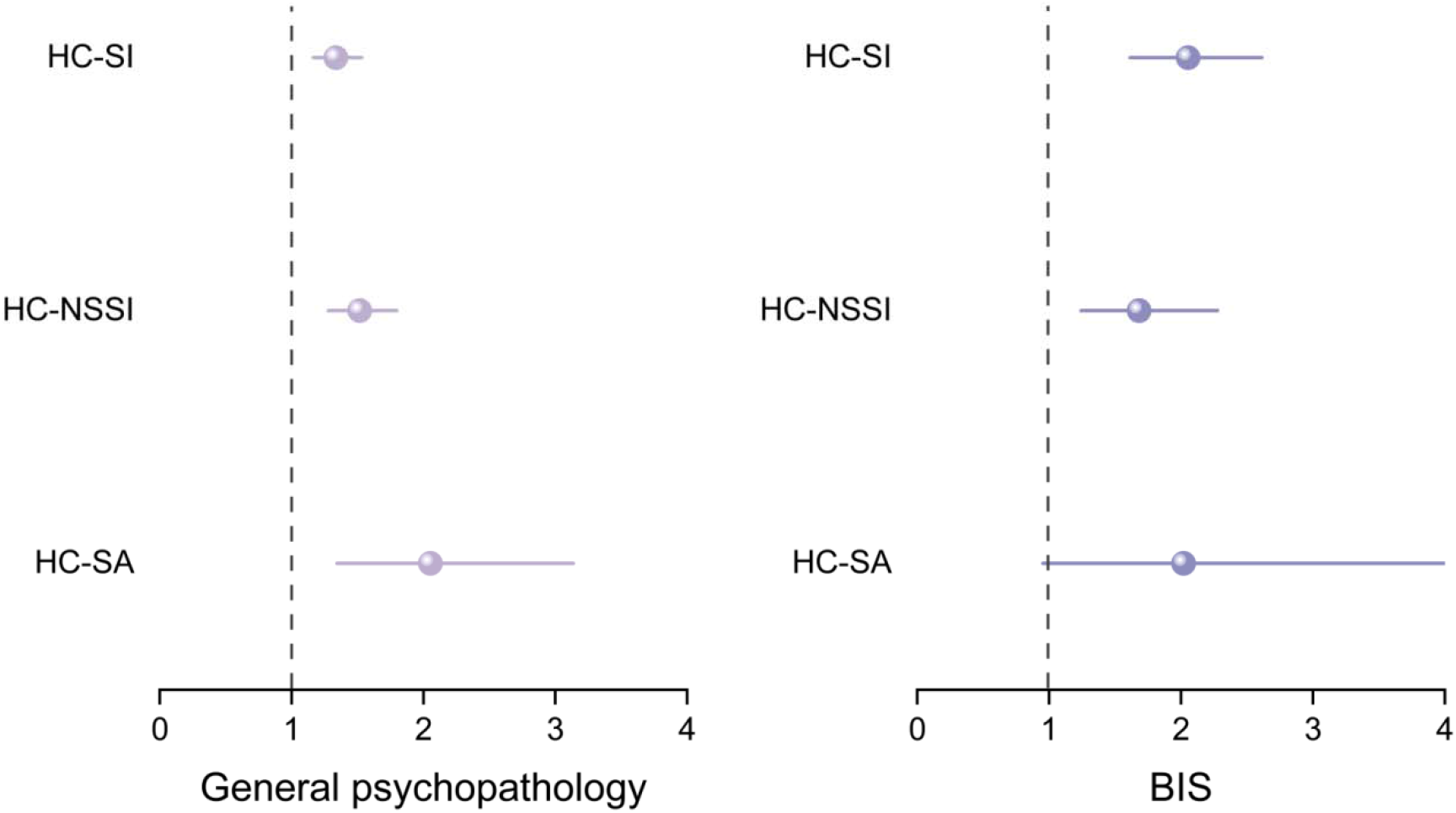
The predictive effect of general psychopathology and BIS on STB and NSSI transitions

### The behavioural system predicts children’s STB and NSSI transition

Group differences in BIS were also found (*F*=20.24, *P*<0.001). In the pairwise comparisons, all the transition groups (i.e., HC-SI, HC-NSSI, and HC-SA) obtained higher BIS scores than the control group (i.e., HC-HC). Besides, as is also shown in Figure 3, a high BIS score predicted high SI (2.05 [1.61, 2.61], and NSSI (1.68 [1.24, 2.28]) transition risk in a year.

### Neurobiological markers of children’s STB and NSSI transition

Overall, abnormalities in the cortical area of the prefrontal cortex (e.g., inferior frontal gyrus; IFG), insula, cingulate cortex, temporal cortex (e.g., inferior temporal sulcus; ITS), precentral cortex, and lateral sulcus were all shown to predict children’s SI, NSSI, and SA transition (all FDR-corrected *P*<0.05). To be more specific, we found that 19 out of the 74 regions could significantly predict children’s SI transition in a year, 15 for NSSI transition, and 29 for the prediction of SA transition (Figure 4). Besides the common cortical regions associated with SI, NSSI, and SA transition simultaneously, SA transition was also predicted by altered cortical areas of the parietal and occipital cortex, suggesting a widely distributed characteristic area.

**Figure 4.**
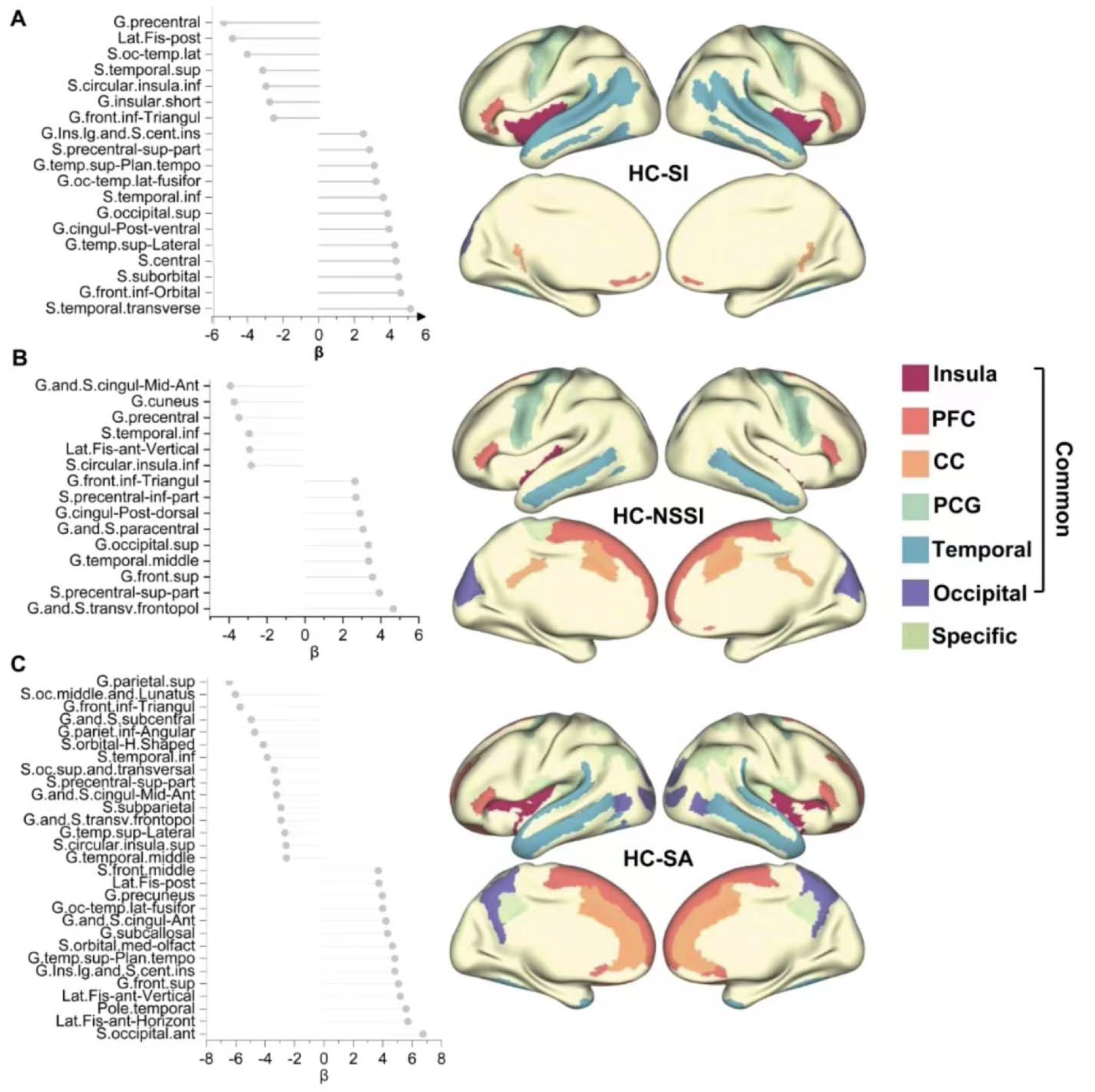
The predictive effect of cortical area on STB and NSSI transitions

## Discussion

Through several findings, this study addressed the gaps in the predictor factors of onset for different subgroups of NSSI or STBs. Our results suggested that 1) the common cortical regions, including the prefrontal cortex (PFC), insula, and cingulate cortex (CC), would predict the onset of NSSI or STBs behaviours; 2) a higher level of *p-factor* was shown to increase the likelihood of adolescents’ persistent of NSSI or STBs behaviours when compared to the healthy group; 3) the *p-factor* also contributes to the development of SA when compared to the NSSI or SI, but no different pathways were found between the occurrence of NSSI and SI; 4) the higher level of impulsivity inhibition abilities (the motivation to avoidance the aversive outcome) was found to increase the likelihood of adolescents’ persistent of NSSI or STBs, but not be a specific risk factor underlying the development of more risky STBs (e.g., SI or SA). Altogether, these findings shed light on the shared and unique risk factors of the occurrence of NSSI or STBs, which would pave the way for the design of future prevention or intervention program to reduce the possibilities of the subsequent mental health crisis.

Potential neurobiological makers for STB and NSSI transition were further detected, including several common cortical regions, including the prefrontal cortex (PFC), insula, and cingulate cortex (CC). These regions’ structural and functional alterations were repeatedly reported to be associated with STB and NSSI in previous studies. ^23,28,29^ Indeed, these brain structure and function disturbances could further lead to alterations in associated brain networks. More specifically, PFC has long been understood as the seat of impulse control,^30^ while high impulsivity was consistently seen as a risk factor for individuals’ STB and NSSI.^30,31^ Therefore, abnormalities in the PFC structure may mediate the associations of impulsivity suppression with STB and NSSI in children. Besides, CC and insula were considered key hubs in pain processing and emotion processing. 11 Moreover, as shown in previous studies, participants with STB or NSSI history always performed aberrant pain perception, ^32,33^and emotion dysregulation.^34,35^ However, the cross-sectional nature of these studies makes it impossible to infer a causal relationship between altered brain structure and individual’s STB and NSSI. Our findings provide novel evidence that the early change in children’s PFC, insula, and CC may serve as neurobiological markers for children’s STB and NSSI transition.

In addition to the individual role of these brain regions, their temporal and spatial associations also significantly impact children’s STB and NSSI transition. For instance, the asynchronous development between PFC and CC would induce children’s risky behaviours,^36^ since CC matures earlier than PFC, which could make children uncontrollably resort to extreme forms of emotional outbursts. Meanwhile, Taylor and colleagues have long identified two resting-state functional connectivity (RSFC) between insula and CC that were closely linked to interception, including the anterior insula to the pregenual anterior cingulate cortex (pACC)/anterior mid-cingulate cortex (aMCC) and the entire insula to mid-cingulate cortex(MCC).^37^ Abnormalities in these RSFCs may lead to interoceptive deficits, which were potential crucial indicators for STB and NSSI risk.^38,39^ Besides, the connection between the insula, CC, and amygdala could potentiate participants’ neuro responses to negative stimuli ^40^, which might also serve as risk factors for children’s latter STB and NSSI.

In line with the previous studies, the *p-factor* as a general factor of psychopathology was functioning as a predictor of individuals at risk for suicide ^17^. However, such findings are generally derived from cross-sectional studies that do not distinguish the temporal sequence of these associations. Our findings extended these previous studies and provided new evidence to our understanding of the underlying mechanism by reporting the *p-factor* could predict the onset of NSSI or STBs longitudinally. Our finding further found that the *p-factor* as a risk factor may distinguish different types of STBs. Specifically, our findings showed that the score of the *p-factor* could be sufficient to distinguish between individuals who develop and persist in NSSI or have SI in a later year and individuals who subsequently engage in suicide attempts ^41^. In line with the ideation-to-action framework, these findings further imply that the score of the *p-factor* may be a high-risk marker for the onset and transition of these serious public health problems. Notwithstanding, the *p-factor* does not show a prominent role in the transition trajectory between the occurrence of NSSI and suicidal ideation. A possible explanation is that the underlying mechanism of NSSI and SI are distinct, with distinct explanations and predictors. For example, NSSI refers to an individual’s self-harm behaviour without the intention to die. More research is needed to explore the potential predictors during these processes further.

Interestingly, we found the prediction role of behavioural inhibition system for the onset of NSSI or STBs overtimes. Given that BIS is sensitive to signals of punishment and concerned with avoiding unfavourable situations ^42^, individuals with a higher tendency of BIS may be more likely to develop self-harm or STBs as the maladaptive coping strategies ^43^. Moreover, the role of the behavioural inhibition system is particularly noteworthy because it has been thought to hasten the transition from suicidal thoughts to action^44^. However, we found that the adolescent’s BIS showed no difference across the NSSI, SI or SA subgroups during the transition processes, which is consistent with a previous study that found that the individual with NSSI showed similarity level of impulsivity when compared to the individual with STBs^45^. Therefore, more research and further corroboration are needed to examine the underlying explanation for the similar level of BIS for different development trajectories of suicide risks.

Nonetheless, several limitations need to be addressed. First, the current studies cannot depict a complete picture of predictor variables for developing various NSSI or STB behaviours among adolescents. Second, the relatively imbalanced sample size for each transition subgroup may lead to measurement bias. Despite that, this study is the first to look at the predictive factors for the different transitions of NSSI and STBs from the biopsychosocial framework. Future research must include more participants in different subgroups to decrease the relatively small and unequal subgroup sizes which may leading to the measurement bias. Second, we only used the two-wave datasets, and the multi-wave longitudinal studies (e.g., ecological momentary assessment) are warranted to capture more dynamic changes during these processes.

Our study provides comprehensive insights into the early signs of suicidality or NSSI transition in children, which cannot be recognized subjectively. The clinical significance of the p-factor, BIS, and different brain regions on individuals’ transition of NSSI or STBs and the changes to high-risk suicide behaviours have warranted our attention. In this case, the early identification of those factors may help to design the early prevention or intervention strategies to prevent the occurrence of more serious public health issues.

## Data Availability

All data produced are available online at https://abcdstudy.org

## Contributors

CRS, ZXQ CZX, WX and QDY designed the study. WX, QDY, LDY and SYN analyses the data. WX, QDY, WYZ, CRS, ZXQ, CZX interpret data. WX and QDY wrote the first draft of the manuscript. WX made the figures. CRS, QDY, CZX, WYZ, ZXQ revised the manuscript critically. All authors contributed feedback and approved the final manuscript.

## Acknowledgments

We thank the Adolescent Brain Cognitive Development (ABCD) participants and their families for their time and dedication to this project. Data used in the preparation of this article were obtained from the ABCD Study (https://abcdstudy.org) and are held in the NIMH Data Archive (NDA). This is a multisite, longitudinal study designed to recruit more than 10,000 children aged 9-10 and follow them over 10 years into early adulthood. The ABCD Study is supported by the National Institutes of Health (NIH)and additional federal partners under award numbers U01DA0401048, U01DA050989, U01DA051016, U01DA041022, U01DA051018, U01DA051037, U01DA050987, U01DA041174, U01DA041106, U01DA041117, U01DA041028, U01DA041134, U01DA050988, U01DA051039, U01DA041156, U01DA041025, U01DA041120, U01DA051038, U01DA041148, U01DA041093, U01DA041089, U24DA041123, U24DA041147. A full list of supporters is available at https://abcdstudy.org/federal-partners.html. A listing of participating sites and a complete listing of the study investigators can be found at https://abcdstudy.org/principal-investigators/. ABCD consortium investigators designed and implemented the study and/or provided data but did not necessarily participate in the analysis or writing of this report. This manuscript reflects the views of the authors and may not reflect the opinions or views of the NIH or ABCD consortium investigators. The ABCD repository grows and changes over time. The ABCD data used in this report came from http://dx.doi.org/10.15154/1523041. DOIs can be found at <u>nda.nih.gov</u>.

